# Why does purpose in life predict mortality in older adults?

**DOI:** 10.1101/2022.03.13.22272312

**Authors:** Richard Sias, H. J. Turtle

## Abstract

**Background:** Previous work documents a strong association between a higher sense of life purpose and lower all-cause mortality risk even when controlling for baseline health and proposes that life purpose intervention may provide a low-cost lever to improve health and longevity. Causation, however, is less clear—lower purpose may cause poorer health and decreased longevity, or poorer health may cause decreased longevity and lower purpose. We examine the extent that (1) more comprehensive health metrics and (2) horizon mitigate or strengthen the relation between purpose and mortality risk to better understand causation.

**Methods:** Prospective cohort sample of 8 425 individuals aged 50 and older who were eligible to participate in the 2006 Health and Retirement Study Psychosocial and Lifestyle questionnaire. Individuals were followed for three subsequent four-year periods: 2006-2010, 2010-2014, and 2014-2018. A total of 1 597 individuals were excluded in the initial four-year period due to lack of follow up, sample weights, or covariates leaving an initial sample of 6 828 individuals. For the second and third four-year periods, an additional 168 and 349 respondents were lost to follow up, respectively. Cox models were estimated to examine the relation between life purpose and mortality for three horizons (years 1-4, 5-8, and 9-12) with more comprehensive measures of current health. Covariates included age, sex, education, race, marital status, smoking status, exercise, alcohol, BMI, and functional health score.

**Findings:** The relation between life purpose and mortality was substantially attenuated or disappeared at longer horizons or when using more comprehensive measures of current health.

**Interpretation:** Much of the documented relation between life purpose and longevity arises from poor health causing higher mortality risk and lower purpose (i.e., reverse causation). As a result, life purpose intervention is likely to be less effective than the previous evidence suggests.

**Funding:** None.

**Research in context:** *Evidence before this study:* We searched PubMed and Google Scholar with no language or date restriction for the term “life purpose” and found four comprehensive reviews of the life purpose or psychological well-being (which included life purpose in the set of psychological well-being metrics) literatures in the last three years and a 2016 meta-analysis of the relation between life purpose and mortality. Although acknowledging it is possible that reverse causation plays a role in linking life purpose levels to subsequent morbidity and mortality, the prevalent view appears to be that even when controlling for current health levels, higher life purpose causes behavioral, biological, or stress buffering changes that, in turn, cause lower future morbidity and mortality.

*Added value of this study:* We demonstrate that the relation between life purpose levels and mortality is substantially attenuated or eliminated when better controlling for current health or focusing on a longer horizon. Both results suggest that the relation between life purpose levels and future mortality risk primarily arises from life purpose proxying for current health levels. The evidence suggests poorer health causes lower life purpose rather than lower life purpose causing poorer health.

*Implications of all the available evidence:* Although life purpose intervention—either at the provider level or in public policy—may have benefits, it is unlikely to cause greater longevity.

## Introduction

Work over the past twenty years demonstrates that a higher sense of life purpose (a measure of eudaimonic well-being) is associated with substantially lower all-cause mortality rates.^1–13^ Given the overwhelming evidence, recent work focuses on understanding how higher purpose *causes* greater longevity via behavioral pathways (e.g., higher purpose might cause healthier eating, greater use of preventive health care, or greater exercise), biological pathways (e.g., greater purpose might cause better lipid profiles, lower blood pressure, or a lesser likelihood of developing diabetes), and psychological stress-buffering pathways (e.g., greater purpose might cause individuals to react less strongly to stressors).^14–24^ As recent studies point out, this evidence is exciting because it represents a potential low-cost lever—life purpose intervention—to improve health and longevity.^1,3,10,14,16,17,25^

Causality, however, is the key remaining question as another potential interpretation of the evidence is that, by definition, poorer baseline health causes increased mortality risk, but could also cause lower life purpose.^14–17,26^ That is, causation may run primarily from health to purpose rather than from purpose to health. Following suggestions in recent surveys,^15,17^ we adopt a two-pronged approach to examining causality by (1) using more comprehensive measures of current health, and (2) evaluating a longer follow-up period.

Most previous work suggests the relation between life purpose and longevity is robust to accounting for current health typically captured by self-rated health,^9,11^ or an indicator for, or count of, chronic disease.^1–3,9,12,27^ Our first approach recognizes that if purpose predicts longevity because poorer baseline health causes lower purpose (i.e., reverse causation), then more comprehensive measures of current health will attenuate the relation between purpose and longevity.

It is also possible, however, that more comprehensive health measures will subsume the relation between purpose and mortality because health and purpose are measured contemporaneously making it difficult to determine whether purpose causes behavioral, biological, or stress-buffering effects that impact mortality risk, or whether health which, by definition causes mortality risk, also causes purpose.^26,28,29^ To address this possibility, our second approach considers how the relation between purpose and mortality changes when examining longer horizons. The intuition is straightforward—if higher purpose causes better health (and therefore longevity), the relation between purpose and longevity should remain, or strengthen, over longer horizons as the causal pathways hypothesized to drive the purpose-longevity relation should impact one’s long-term mortality risk as much as they impact one’s near-term mortality risk. For example, evidence suggests smoking, hypertension, lung function, diet, exercise, lipid profiles, and preventive health care impact long-term outcomes as much as they impact near-term outcomes.^30–33^ That is, healthier eating (a behavioral pathway), healthier lipid profiles (a biological pathway), or better dealing with stress (a stress-buffering pathway) should impact long-term longevity at least as much as near-term longevity as the effects are often cumulative. Moreover, the life purpose literature suggests purpose generates a virtuous cycle—higher purpose begets better health which leads to even higher purpose implying the relation between purpose and mortality should strengthen with horizon.^34^

In contrast, if purpose predicts mortality because current health levels impact purpose (i.e., reverse causality), then the relation between purpose and mortality will attenuate with time as those who are ill today (i.e., poor current health) will drop from the sample as they die. This idea is widely used in the literature as many studies either exclude individuals with existing serious medical conditions (e.g., those diagnosed with cancer) or those who die in the first year of follow up.^1,2,11,12,27^ Importantly, however, given (1) most serious illnesses have survival rates much greater than one year,^35,36^ and (2) many ill individuals remain undiagnosed,^37^ a one-year exclusion or the exclusion of individuals diagnosed with a set of specific diseases is unlikely to eliminate most seriously ill respondents from the sample. We extend this line of reasoning—if life purpose predicts longevity because a decline in health causes a decline in purpose, then the relation between purpose and longevity should be somewhat attenuated when eliminating those individuals who die in the first year, more attenuated when excluding those who die in the first four years, and even more attenuated when excluding those who die in the first eight years. In sum, if the purpose-mortality relation primarily arises because purpose causes better health and longevity then the relation between purpose and longevity should be as strong or stronger with longer horizons. In contrast, if the relation primarily arises because health causes purpose, then the relation between purpose and longevity will weaken with horizon.

## Methods

### Study design and participants

Our strategy is to replicate a recent well-executed study of the relation between purpose and longevity and then examine the sensitivity of the results to horizon and more comprehensive measures of current health. Specifically, we use the Health and Retirement Study data (HRS is sponsored by the National Institute on Aging, grant number U01AG009740, and is conducted by the University of Michigan) to replicate and extend the Alimujiang et al. study—arguably the most comprehensive to date as it uses both the broadest set of controls (including “sociodemographic characteristics, baseline health and behavioral characteristics, and psychological factors”) and the most validated life purpose measure.^1,38^ The authors find respondents within the lowest life purpose group are 215% (i.e., HR=3·15, *p*<0·001) more likely to die in the subsequent four years than respondents in the highest life purpose group when controlling for standard covariates and 143% (i.e., HR=2·43, *p*<0·001) more likely to die when additionally controlling for psychological constructs.

### Procedures

HRS is a nationally representative biennial prospective cohort survey of individuals age 50 and older and their spouses.^39,40^ In 2006, HRS randomly selected half the participants for an enhanced face-to-face interview and a leave-behind Psychosocial and Lifestyle questionnaire that included an assessment of life purpose. We use the psychosocial and lifestyle sample weights that adjust for both core sample interview weights and psychosocial sample non-response.^41^ As detailed in the appendix (pp 1-5), the initial sample consists of 8 425 individuals age 50 and older who were eligible to complete the questionnaire of which 7 282 had life purpose data. Another 454 individuals were dropped from the sample due to missing covariates or weights leaving an initial sample of 6 828 individuals for the 2006-2010 period. Another 168 and 349 respondents are lost due to missing follow up data for the 2010-2014 and 2014-2018 periods, respectively. Because the study uses deidentified publicly available information, it was exempted from University of Arizona IRB review.

HRS identifies death dates via next of kin interviews (and confirms with the National Death Index).^42^ Of the 6 828 individuals in our sample, 765 died (11·2%) within the initial four years (2006-2010). We censored those who were interviewed in 2010 at their 2010 interview date (n=5 821). For the 158 respondents who did not complete a 2010 interview but were reported alive or died in a subsequent interview wave, we censored at 48 months beyond their 2006 interview date. For the 84 individuals who completed a 2008 interview but did not complete any subsequent interview and were not reported as having died in any subsequent wave, we censored at the 2008 interview date (appendix, p 3).

The second four-year period (2010-2014) sample consists of 5 895 individuals. A total of 824 respondents died during the 2010-2014 period (14·0%). Survivors include 4 813 individuals interviewed for the 2014 wave (censored at their 2014 interview date), 133 respondents who did not complete a 2014 interview but either completed a subsequent (2016 or 2018) interview or were reported to have died in a subsequent wave (censored at 96 months following their 2006 interview; see appendix p. 4), and 125 individuals last interviewed in 2012 (censored at their 2012 interview date).

The final four-year period (2014-2018) consists of 4 722 respondents. A total of 871 individuals died during this period of whom 782 had reported death dates and 89 were reported to have died between the 2016 and 2018 interviews but death date was unavailable (we assign these 89 individuals a death date of July 2017). The surviving sample consists of 3 433 individuals who were interviewed in 2018 and assigned a censored date of the 2018 interview and 418 individuals last interviewed in 2016 and assigned a censored date of their 2016 interview (appendix p 5).

### Outcomes and covariates

Life purpose is assessed by the Ryff and Keyes seven-question measure.^38^ Individuals respond to each statement (e.g., “I enjoy making plans for the future and working to make them a reality”) on a six-point Likert scale ranging from “1=Strongly disagree” to “6=Strongly agree.” Following HRS guidance,^41^ we reverse score the four “negative” statements and then average the score across all seven statements for respondents who answer at least four statements. Replicating Alimujiang et al.,^1^ we partition respondents into five groups by life purpose score—those between 1·00-2·99 (lowest life purpose group), 3·00-3·99, 4·00-4·99, 5·00-5·99, and 6·0.

Following Alimujiang et al.,^1^ the initial model includes four health behaviors (smoking status, alcohol consumption, vigorous physical activities, and BMI) and two current health metrics—an indicator for the presence of one or more chronic diseases (hypertension, diabetes, cancer, lung disease, heart disease, or stroke) and a functional status score that ranges from 0 to 5 and is the summation of whether the individual responds yes to questions of whether they have “some difficulty” (1) walking across a room, (2) bathing, (3) eating, (4) dressing, and (5) getting in and out of bed.

We include the four health behaviors in all subsequent models but consider three more comprehensive measures of current health. First, we replace the chronic disease indicator with indicators for each of the six chronic diseases (e.g., hypertension) as the hazard ratios vary dramatically across the six chronic diseases (see appendix p 11). Second, we include the respondents’ self-rated health. Specifically, respondents are asked “Would you say your health is excellent (1), very good (2), good (3), fair (4) or poor (5)?” We reverse score the variable so that higher values represent better health. Third, we construct a more objective (relative to self-rated health) measure of current health, that we denote “broad limitations,” as the first principal component from the four RAND-calculated functional limitations metrics based on whether respondents have some difficulty with Mobility, Large Muscles, Gross Motor Skills, and Fine Motor Skills (see appendix p 12).^43^ Additional covariates include respondent sex, age, education, and marital status. For the authors’ Model 2, we also compute seven psychological constructs—depression, anxiety, cynical hostility, negative affect, optimism, positive affect, and social participation.^1^ As detailed in the appendix (p 2), including the psychological constructs reduces the sample size by approximately 7%. See appendix (pp 9-10) for construction details for all variables.

### Statistical analysis

We computed descriptive statistics for all variables. Cox proportional hazard models were used to replicate the previous study examining the relation between 2006 life purpose and survival over the 2006-2010 period.^1^ We then examined how the hazard rates were impacted when (1) using more comprehensive health metrics, and (2) examining survival over subsequent periods. All analyses were conducted from October 10, 2021 to February 28, 2022, using SAS version 9·4. A two-tailed *p*<0·05 was considered statistically significant.

## Results

Table 1 reports descriptive statistics (all variables are measured in the 2006 interview) for the three four-year periods. By construction, the results for the initial period are nearly identical to those reported by the authors (appendix p 6).^1^ Because purpose and self-reported health both decline with age (appendix p 7) and mortality risk rises with age, the surviving sample’s 2006 life purpose, self-rated health, and mortality rates rise, and the average (2006) age declines, over the three periods.

**Table 1.** Descriptive characteristics of 2006 Health and Retirement Study (HRS) participants for three successive 4-y periods.

| Characteristic | Participants, No. (%)<br>2006-2010 |  | Participants, No. (%)<br>2010-2014 |  | Participants, No. (%)<br>2014-2018 |  |
| --- | --- | --- | --- | --- | --- | --- |
|  | No event<br>(n=6 063) | Death<br>(n=765) | No event<br>(n=5 071) | Death<br>(n=824) | No event<br>(n=3 851) | Death<br>(n=871) |
| Age, y |  |  |  |  |  |  |
| 50-54 | 471 (7·8) | 13 (1·7) | 435 (8·6) | 20 (2·4) | 388 (10·1) | 15 (1·7) |
| 55-59 | 971 (16) | 36 (4·7) | 911 (18) | 35 (4·2) | 791 (20·5) | 49 (5·6) |
| 60-64 | 938 (15·5) | 54 (7·1) | 857 (16·9) | 55 (6·7) | 735 (19·1) | 74 (8·5) |
| 65-69 | 1 246 (20·6) | 117 (15·3) | 1 085 (21·4) | 119 (14·4) | 861 (22·4) | 151 (17·3) |
| 70-74 | 1 030 (17) | 118 (15·4) | 862 (17) | 143 (17·4) | 616 (16) | 178 (20·4) |
| 75-79 | 707 (11·7) | 128 (16·7) | 531 (10·5) | 157 (19·1) | 318 (8·3) | 181 (20·8) |
| 80+ | 700 (11·5) | 299 (39·1) | 390 (7·7) | 295 (35·8) | 142 (3·7) | 223 (25·6) |
| Sex |  |  |  |  |  |  |
| Male | 2 539 (41·9) | 376 (49·2) | 2 052 (40·5) | 418 (50·7) | 1 524 (39·6) | 388 (44·5) |
| Female | 3 524 (58·1) | 389 (50·8) | 3 019 (59·5) | 406 (49·3) | 2 327 (60·4) | 483 (55·5) |
| Marital Status |  |  |  |  |  |  |
| Married | 4 203 (69·3) | 412 (53·9) | 3 589 (70·8) | 493 (59·8) | 2 798 (72·7) | 532 (61·1) |
| Separated or divorced | 664 (11) | 65 (8·5) | 569 (11·2) | 74 (9) | 444 (11·5) | 82 (9·4) |
| Widowed | 1 033 (17) | 258 (33·7) | 772 (15·2) | 243 (29·5) | 496 (12·9) | 237 (27·2) |
| Never married | 163 (2·7) | 30 (3·9) | 141 (2·8) | 14 (1·7) | 113 (2·9) | 20 (2·3) |
| Race/ethnicity |  |  |  |  |  |  |
| Non-Hispanic white | 4 745 (78·3) | 618 (80·8) | 3 949 (77·9) | 656 (79·6) | 2 967 (77) | 701 (80·5) |
| Non-Hispanic and<br>Hispanic black | 762 (12·6) | 93 (12·2) | 637 (12·6) | 114 (13·8) | 487 (12·6) | 123 (14·1) |
| Hispanic white | 307 (5·1) | 38 (5) | 268 (5·3) | 34 (4·1) | 219 (5·7) | 28 (3·2) |
| Other | 249 (4·1) | 16 (2·1) | 217 (4·3) | 20 (2·4) | 178 (4·6) | 19 (2·2) |
| Educational level |  |  |  |  |  |  |
| <High school | 1 017 (16·8) | 230 (30·1) | 803 (15·8) | 198 (24) | 548 (14·2) | 214 (24·6) |
| High school | 2 254 (37·2) | 282 (36·9) | 1 863 (36·7) | 325 (39·4) | 1 391 (36·1) | 329 (37·8) |
| Some college | 1 391 (22·9) | 147 (19·2) | 1 187 (23·4) | 171 (20·8) | 928 (24·1) | 176 (20·2) |
| College | 837 (13·8) | 59 (7·7) | 718 (14·2) | 86 (10·4) | 570 (14·8) | 99 (11·4) |
| Graduate school | 564 (9·3) | 47 (6·1) | 500 (9·9) | 44 (5·3) | 414 (10·8) | 53 (6·1) |
| Smoking Status |  |  |  |  |  |  |
| Never | 2 690 (44·4) | 249 (32·5) | 2 321 (45·8) | 292 (35·4) | 1 794 (46·6) | 365 (41·9) |
| Current smoker | 764 (12·6) | 122 (15·9) | 620 (12·2) | 120 (14·6) | 458 (11·9) | 123 (14·1) |
| Former smoker | 2 609 (43) | 394 (51·5) | 2 130 (42) | 412 (50) | 1 599 (41·5) | 383 (44) |
| Alcohol, days drink/wk |  |  |  |  |  |  |
| 0 | 3 959 (65·3) | 574 (75) | 3 282 (64·7) | 581 (70·5) | 2 445 (63·5) | 617 (70·8) |
| 1-2 | 991 (16·3) | 101 (13·2) | 862 (17) | 102 (12·4) | 695 (18) | 106 (12·2) |
| 3-4 | 402 (6·6) | 23 (3) | 350 (6·9) | 37 (4·5) | 269 (7) | 49 (5·6) |
| 5-6 | 204 (3·4) | 13 (1·7) | 179 (3·5) | 19 (2·3) | 144 (3·7) | 23 (2·6) |
| Every day | 507 (8·4) | 54 (7·1) | 398 (7·8) | 85 (10·3) | 298 (7·7) | 76 (8·7) |
| Vigorous phy. exercise |  |  |  |  |  |  |
| Every day | 171 (2·8) | 21 (2·7) | 149 (2·9) | 16 (1·9) | 113 (2·9) | 22 (2·5) |
| More than once a week | 1317 (21·7) | 78 (10·2) | 1 155 (22·8) | 118 (14·3) | 927 (24·1) | 148 (17) |
| Once a week | 493 (8·1) | 32 (4·2) | 431 (8·5) | 46 (5·6) | 354 (9·2) | 54 (6·2) |
| 1-3 Times a month | 416 (6·9) | 25 (3·3) | 372 (7·3) | 32 (3·9) | 303 (7·9) | 43 (4·9) |
| Hardly ever or never | 3 666 (60·5) | 609 (79·6) | 2 964 (58·5) | 612 (74·3) | 2 154 (55·9) | 604 (69·3) |
| Chronic illness |  |  |  |  |  |  |
| No | 1 662 (27·4) | 77 (10·1) | 1 492 (29·4) | 105 (12·7) | 1 235 (32·1) | 145 (16·6) |
| Yes | 4 401 (72·6) | 688 (89·9) | 3 579 (70·6) | 719 (87·3) | 2 616 (67·9) | 726 (83·4) |
|  | No event<br>(n=6063) | Death<br>(n=765) | No event<br>(n=5071) | Death<br>(n=824) | No event<br>(n=3851) | Death<br>(n=871) |
| Body mass index |  |  |  |  |  |  |
| ≤ 18·50 | 57 (0·9) | 38 (5) | 43 (0·8) | 13 (1·6) | 28 (0·7) | 13 (1·5) |
| 18·51-24·99 | 1 666 (27·5) | 278 (36·3) | 1 343 (26·5) | 264 (32) | 974 (25·3) | 273 (31·3) |
| 25·00-29·99 | 2 381 (39·3) | 239 (31·2) | 2 000 (39·4) | 312 (37·9) | 1 534 (39·8) | 309 (35·5) |
| ≥ 30·00 | 1 959 (32·3) | 210 (27·5) | 1 685 (33·2) | 235 (28·5) | 1 315 (34·1) | 276 (31·7) |
| Life purpose category |  |  |  |  |  |  |
| 1·00-2·99 | 204 (3·4) | 73 (9·5) | 155 (3·1) | 41 (5) | 113 (2·9) | 35 (4) |
| 3·00-3·99 | 1 352 (22·3) | 275 (35·9) | 1 033 (20·4) | 272 (33) | 722 (18·7) | 252 (28·9) |
| 4·00-4·99 | 2 148 (35·4) | 265 (34·6) | 1 821 (35·9) | 279 (33·9) | 1 387 (36) | 305 (35) |
| 5·00-5·99 | 1 961 (32·3) | 132 (17·3) | 1 704 (33·6) | 206 (25) | 1 342 (34·8) | 235 (27) |
| 6·00 | 398 (6·6) | 20 (2·6) | 358 (7·1) | 26 (3·2) | 287 (7·5) | 44 (5·1) |
| High blood pressure |  |  |  |  |  |  |
| No | 2 636 (43·5) | 253 (33·1) | 2 269 (44·7) | 281 (34·1) | 1 805 (46·9) | 301 (34·6) |
| Yes | 3 427 (56·5) | 512 (66·9) | 2 802 (55·3) | 543 (65·9) | 2 046 (53·1) | 570 (65·4) |
| Diabetes |  |  |  |  |  |  |
| No | 4 918 (81·1) | 533 (69·7) | 4 163 (82·1) | 602 (73·1) | 3 211 (83·4) | 658 (75·5) |
| Yes | 1 145 (18·9) | 232 (30·3) | 908 (17·9) | 222 (26·9) | 640 (16·6) | 213 (24·5) |
| Cancer |  |  |  |  |  |  |
| No | 5 212 (86) | 562 (73·5) | 4 431 (87·4) | 634 (76·9) | 3 430 (89·1) | 690 (79·2) |
| Yes | 851 (14) | 203 (26·5) | 640 (12·6) | 190 (23·1) | 421 (10·9) | 181 (20·8) |
| Lung disease |  |  |  |  |  |  |
| No | 5 520 (91) | 601 (78·6) | 4 673 (92·2) | 686 (83·3) | 3 581 (93) | 768 (88·2) |
| Yes | 543 (9) | 164 (21·4) | 398 (7·8) | 138 (16·7) | 270 (7) | 103 (11·8) |
| Heart disease |  |  |  |  |  |  |
| No | 4 684 (77·3) | 412 (53·9) | 4 045 (79·8) | 499 (60·6) | 3 193 (82·9) | 562 (64·5) |
| Yes | 1 379 (22·7) | 353 (46·1) | 1 026 (20·2) | 325 (39·4) | 658 (17·1) | 309 (35·5) |
| Stroke |  |  |  |  |  |  |
| No | 5 760 (95) | 672 (87·8) | 4 852 (95·7) | 746 (90·5) | 3 722 (96·7) | 789 (90·6) |
| Yes | 303 (5) | 93 (12·2) | 219 (4·3) | 78 (9·5) | 129 (3·3) | 82 (9·4) |
| Continuous variables: Mean (SD) |  |  |  |  |  |  |
| Functional score | 0·22 (0·68) | 0·76 (1·26) | 0·18 (0·61) | 0·46 (0·99) | 0·16 (0·57) | 0·29 (0·76) |
| Life purpose score | 4·58 (0·91) | 4·09 (0·95) | 4·63 (0·9) | 4·31 (0·92) | 4·67 (0·89) | 4·41 (0·9) |
| Self-rated health | 3·29 (1·06) | 2·48 (1·08) | 3·36 (1·04) | 2·83 (1·07) | 3·44 (1·02) | 2·98 (1·02) |
| Broad limitations | -0·09 (0·92) | 0·71 (1·31) | -0·16 (0·86) | 0·36 (1·15) | -0·21 (0·81) | 0·12 (0·99) |

Number of respondents in each variable response category for all variables within each HRS wave (2006-2010, 2010-2014, 2014-2018). Percent responses by variable-wave response are provided for each variable in parentheses.

As detailed in the appendix (p 13), tests reveal the constant proportional hazards assumption is violated over the 12-year period as the relation between purpose and mortality declines with horizon. Thus, Table 2 reports Cox models for three different periods. Model 1 is identical to Alimujiang et al.’s Model 1 and includes controls for age, sex, education, marital status, smoking status, frequency of vigorous physical activity, days/week consuming alcohol, BMI, functional status, and an indicator for the presence of a chronic condition.^1^ Model 1A makes two changes (1) replacing the indicator for any chronic disease with indicators for each of the six chronic conditions and (2) adding self-rated health as a covariate. Model 1B differs from Model 1A by replacing the functional score and self-rated health score with the broad limitations index.

**Table 2.**
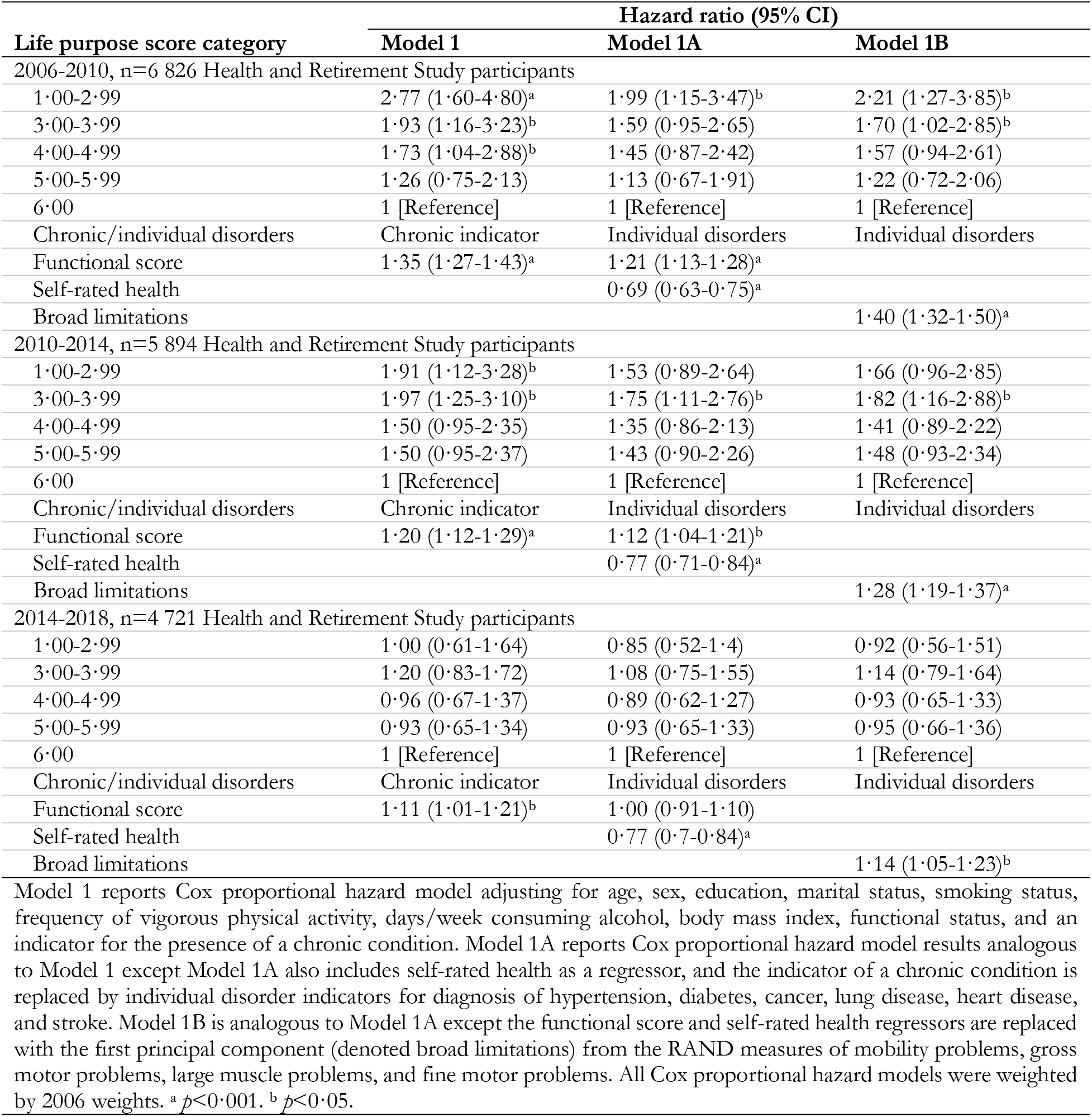
Life purpose, health, and all-cause mortality.

| Life purpose score category | Hazard ratio (95% CI) |  |  |
| --- | --- | --- | --- |
|  | Model 1 | Model 1A | Model 1B |
| 2006-2010, n=6 826 Health and Retirement Study participants |  |  |  |
| 1·00-2·99 | 2·77 (1·60-4·80)<sup>a</sup> | 1·99 (1·15-3·47)<sup>b</sup> | 2·21 (1·27-3·85)<sup>b</sup> |
| 3·00-3·99 | 1·93 (1·16-3·23)<sup>b</sup> | 1·59 (0·95-2·65) | 1·70 (1·02-2·85)<sup>b</sup> |
| 4·00-4·99 | 1·73 (1·04-2·88)<sup>b</sup> | 1·45 (0·87-2·42) | 1·57 (0·94-2·61) |
| 5·00-5·99 | 1·26 (0·75-2·13) | 1·13 (0·67-1·91) | 1·22 (0·72-2·06) |
| 6·00 | 1 [Reference] | 1 [Reference] | 1 [Reference] |
| Chronic/individual disorders | Chronic indicator | Individual disorders | Individual disorders |
| Functional score | 1·35 (1·27-1·43)<sup>a</sup> | 1·21 (1·13-1·28)<sup>a</sup> |  |
| Self-rated health |  | 0·69 (0·63-0·75)<sup>a</sup> |  |
| Broad limitations |  |  | 1·40 (1·32-1·50)<sup>a</sup> |
| 2010-2014, n=5 894 Health and Retirement Study participants |  |  |  |
| 1·00-2·99 | 1·91 (1·12-3·28)<sup>b</sup> | 1·53 (0·89-2·64) | 1·66 (0·96-2·85) |
| 3·00-3·99 | 1·97 (1·25-3·10)<sup>b</sup> | 1·75 (1·11-2·76)<sup>b</sup> | 1·82 (1·16-2·88)<sup>b</sup> |
| 4·00-4·99 | 1·50 (0·95-2·35) | 1·35 (0·86-2·13) | 1·41 (0·89-2·22) |
| 5·00-5·99 | 1·50 (0·95-2·37) | 1·43 (0·90-2·26) | 1·48 (0·93-2·34) |
| 6·00 | 1 [Reference] | 1 [Reference] | 1 [Reference] |
| Chronic/individual disorders | Chronic indicator | Individual disorders | Individual disorders |
| Functional score | 1·20 (1·12-1·29)<sup>a</sup> | 1·12 (1·04-1·21)<sup>b</sup> |  |
| Self-rated health |  | 0·77 (0·71-0·84)<sup>a</sup> |  |
| Broad limitations |  |  | 1·28 (1·19-1·37)<sup>a</sup> |
| 2014-2018, n=4 721 Health and Retirement Study participants |  |  |  |
| 1·00-2·99 | 1·00 (0·61-1·64) | 0·85 (0·52-1·4) | 0·92 (0·56-1·51) |
| 3·00-3·99 | 1·20 (0·83-1·72) | 1·08 (0·75-1·55) | 1·14 (0·79-1·64) |
| 4·00-4·99 | 0·96 (0·67-1·37) | 0·89 (0·62-1·27) | 0·93 (0·65-1·33) |
| 5·00-5·99 | 0·93 (0·65-1·34) | 0·93 (0·65-1·33) | 0·95 (0·66-1·36) |
| 6·00 | 1 [Reference] | 1 [Reference] | 1 [Reference] |
| Chronic/individual disorders | Chronic indicator | Individual disorders | Individual disorders |
| Functional score | 1·11 (1·01-1·21)<sup>b</sup> | 1·00 (0·91-1·10) |  |
| Self-rated health |  | 0·77 (0·7-0·84)<sup>a</sup> |  |
| Broad limitations |  |  | 1·14 (1·05-1·23)<sup>b</sup> |

Model 1 reports Cox proportional hazard model adjusting for age, sex, education, marital status, smoking status, frequency of vigorous physical activity, days/week consuming alcohol, body mass index, functional status, and an indicator for the presence of a chronic condition. Model 1A reports Cox proportional hazard model results analogous to Model 1 except Model 1A also includes self-rated health as a regressor, and the indicator of a chronic condition is replaced by individual disorder indicators for diagnosis of hypertension, diabetes, cancer, lung disease, heart disease, and stroke. Model 1B is analogous to Model 1A except the functional score and self-rated health regressors are replaced with the first principal component (denoted broad limitations) from the RAND measures of mobility problems, gross motor problems, large muscle problems, and fine motor problems. All Cox proportional hazard models were weighted by 2006 weights. <sup>a</sup> $p < 0·001$ . <sup>b</sup> $p < 0·05$ .

Although hazard ratios are slightly smaller, the results for Model 1 are consistent with Alimujiang et al.,^1^ and indicate that individuals with the lowest life purpose are 177% more likely to die within the first four years than individuals with the highest purpose. The relation between purpose and longevity, however, quickly deteriorates with horizon as individuals with low purpose are 91% more likely to die (under Model 1) than those with the high purpose over the subsequent four years (2010-2014), i.e., the relative risk is reduced by approximately half (i.e., 1-(1.91-1)/(2.77-1)) when eliminating individuals who die in the initial four-year period. Over years 8-12 (the 2014-2018 period), there is no evidence those with the lowest purpose have greater mortality risk than others (HR, 1·00: 95% CI, 0·61-1·64).

Model 1A reveals that adding self-rated health and indicators for individual chronic conditions also substantially attenuates the purpose-longevity relation. Over the initial four-year period, these two changes result in a 44% reduction in excess risk of low life purpose individuals (i.e., 1-(1·99-1)/(2·77-1)). Similarly, relative to the baseline model, the inclusion of these variables results in a 70% reduction (i.e., 1-(1·53-1)/(2·77-1)) in the relative risk of low purpose individuals in the second four-year period (2010-2014) and the relation between the purpose categories and mortality becomes non-monotonic. Although 2006 self-rated health continues to strongly predict longevity in years 8-12, there is no evidence that purpose is related to longevity over the final four-year period (2014-2018) as the point estimate for the hazard ratio drops below one.

Some suggest that controlling for self-rated health may be an “overadjustment” because self-rated health,^17^ “…is both defined and influenced by functional health, physical conditions, and, most importantly, psychological distress and well-being…”^29,44^ Thus, Model 1B replaces self-rated health with a more objective health metric—the broad limitations index. The result in the final column reveal that the broad limitations metric also substantially attenuates the relation between purpose and longevity as the excess risk of low purpose respondents is reduced by 32% relative to the baseline Model 1 (i.e., 1-(2·21-1)/(2·77-1)) in the initial four-year period, reduced by 63% (i.e., 1-(1·66-1)/(2·77-1)) in the next four years, and reduced by 84% (i.e., 1-(1·28-1)/(2·77-1)) in the final four-year period. Moreover, although there is no evidence that those in the highest purpose group have lower mortality risk than those in the lowest group beyond the initial four-year period, the broad limitations metric continues to forecast longevity over both years 5-8 and 9-12.

The first column of Table 3 reports Alimujiang et al.’s Model 2 that adds the seven psychological constructs.^1^ Consistent with their evidence, the psychological constructs attenuate the relation between purpose and longevity. As noted by the authors, the attenuation suggests that the relation between purpose and longevity is either mediated or confounded by these psychological constructs. Regardless, after including the psychological constructs, there is no evidence that individuals with low purpose have greater mortality risk than those with high purpose beyond the initial four-year period. For instance, the relative risk falls 81% (i.e., 1-(1·23-1)/(2·18-1)) in the subsequent four-year period relative to the initial four-year period. Moreover, although all the health metrics (functional score, self-rated health, and broad limitations) continue to predict mortality risk in the subsequent four-year period (and self-rated health continues to predict mortality in the final four-year period), there is no evidence purpose is related to mortality in any of the six models (2, 2A, or 2B) for the subsequent, or final, four-year period.

**Table 3.** Life purpose, health, psychological constructs, and all-cause mortality.

| Life purpose score category | Hazard Ratio (95% CI) |  |  |
| --- | --- | --- | --- |
|  | Model 2 | Model 2A | Model 2B |
| 2006-2010, n=6 356 Health and Retirement Study participants |  |  |  |
| 1·00-2·99 | 2·18 (1·20-3·95)<sup>b</sup> | 1·89 (1·04-3·44)<sup>b</sup> | 1·86 (1·02-3·38)<sup>b</sup> |
| 3·00-3·99 | 1·64 (0·95-2·81) | 1·57 (0·92-2·70) | 1·54 (0·89-2·63) |
| 4·00-4·99 | 1·46 (0·86-2·48) | 1·38 (0·81-2·33) | 1·37 (0·81-2·32) |
| 5·00-5·99 | 1·15 (0·67-1·96) | 1·10 (0·64-1·88) | 1·12 (0·66-1·91) |
| 6·00 | 1 [Reference] | 1 [Reference] | 1 [Reference] |
| Chronic/individual disorders | Chronic indicator | Individual disorders | Individual disorders |
| Functional score | 1·33 (1·24-1·43)<sup>a</sup> | 1·21 (1·12-1·3)<sup>a</sup> |  |
| Self-rated health |  | 0·70 (0·63-0·77)<sup>a</sup> |  |
| Broad limitations |  |  | 1·40 (1·30-1·51)<sup>a</sup> |
| 2010-2014, n=5 525 Health and Retirement Study participants |  |  |  |
| 1·00-2·99 | 1·23 (0·67-2·25) | 1·16 (0·63-2·12) | 1·14 (0·62-2·10) |
| 3·00-3·99 | 1·47 (0·89-2·42) | 1·47 (0·89-2·42) | 1·44 (0·87-2·38) |
| 4·00-4·99 | 1·23 (0·76-2·00) | 1·19 (0·73-1·94) | 1·20 (0·74-1·95) |
| 5·00-5·99 | 1·44 (0·89-2·34) | 1·41 (0·87-2·29) | 1·42 (0·88-2·31) |
| 6·00 | 1 [Reference] | 1 [Reference] | 1 [Reference] |
| Chronic/individual disorders | Chronic indicator | Individual disorders | Individual disorders |
| Functional score | 1·18 (1·09-1·28)<sup>a</sup> | 1·11 (1·02-1·21)<sup>b</sup> |  |
| Self-rated health |  | 0·74 (0·68-0·82)<sup>a</sup> |  |
| Broad limitations |  |  | 1·25 (1·16-1·36)<sup>a</sup> |
| 2014-2018, n=4 450 Health and Retirement Study participants |  |  |  |
| 1·00-2·99 | 0·71 (0·41-1·22) | 0·67 (0·39-1·16) | 0·66 (0·38-1·14) |
| 3·00-3·99 | 0·91 (0·61-1·36) | 0·88 (0·59-1·32) | 0·88 (0·59-1·31) |
| 4·00-4·99 | 0·77 (0·53-1·13) | 0·75 (0·51-1·09) | 0·75 (0·52-1·10) |
| 5·00-5·99 | 0·86 (0·59-1·25) | 0·86 (0·59-1·25) | 0·86 (0·59-1·25) |
| 6·00 | 1 [Reference] | 1 [Reference] | 1 [Reference] |
| Chronic/individual disorders | Chronic indicator | Individual disorders | Individual disorders |
| Functional score | 1·04 (0·94-1·15) | 0·95 (0·86-1·06) |  |
| Self-rated health |  | 0·78 (0·71-0·86)<sup>a</sup> |  |
| Broad limitations |  |  | 1·08 (0·98-1·18) |

Model 2 reports results for the Cox proportional hazard model adjusting for age, sex, education, marital status, smoking status, frequency of vigorous physical activity, days/week consuming alcohol, body mass index, functional status, seven psychological constructs (depression, anxiety, cynical hostility, negative affect, optimism, positive affect, and social participation), and an indicator for the presence of a chronic condition. Model 2A reports Cox proportional hazard model results analogous to Model 2 except Model 2A also includes self-rated health as a regressor, and the indicator of a chronic condition is replaced by individual disorder indicators for diagnosis of hypertension, diabetes, cancer, lung disease, heart disease, and stroke. Model 2B is analogous to Model 2A except the functional score and self-rated health regressors are replaced with the first principal component (denoted broad limitations) from the RAND measures of mobility problems, gross motor problems, large muscle problems, and fine motor problems. All Cox proportional hazard models were weighted by 2006 weights. <sup>a</sup> $p < 0·001$ . <sup>b</sup> $p < 0·05$ .

## Discussion

Consistent with previous work, higher life purpose was inversely related to mortality risk over the subsequent four-year period. Relative to the baseline model, the higher mortality risk of low life purpose respondents fell by nearly 50% in the subsequent four-year period and by 100% in years 8-12. Including more comprehensive current health metrics also substantially attenuated the relative risks. Regardless of the model, we found no evidence that purpose was meaningfully related to mortality risk beyond the initial eight-year period when excluding the psychological constructs or beyond the initial four-year period when including the psychological constructs.

The substantial attenuation of the purpose-mortality relation when using more comprehensive health measures suggests that either health drives purpose (i.e., reverse causation) or that purpose predicts longevity because purpose causes current health levels. The attenuation or elimination of the relation between purpose and longevity at longer horizons provides further evidence for the reverse causation explanation. That is, at longer horizons (e.g., years 8-12), many individuals suffering from poor health (diagnosed or undiagnosed) in 2006 will have died and the remaining 2014-2018 sample consists of individuals in relatively good health in 2006 but who vary with respect to life purpose in 2006 and whose 2006 life purpose is unaffected by post-2006 changes in health (i.e., relatively free of reverse causation).

One concern is that the smaller sample available in the final period means the tests are underpowered to identify the relation between purpose and longevity (although the 2006 health metrics continue to predict long-term survival). Inconsistent with this explanation, however, when the sample is limited to individuals with both 2006 and 2014 data, 2014 purpose is strongly related to 2014-2018 mortality (HR, 1·94: 95% CI, 1·23-3·07), but 2006 purpose is not (HR, 0·90: 95% CI, 0·48-1·69, see appendix pp 14-16). This means that either changes in health substantially impact purpose (reverse causation), or changes in purpose have strong near-term health impacts but purpose levels do not impact mortality risk. That is, a large decline in 2006-2014 purpose increases mortality risk, but those who had low life purpose in both 2006 and 2014 do not face increased mortality risk. The latter interpretation is hard to reconcile with the hypothesized causal pathways between purpose and longevity, e.g., that low purpose causes unhealthier eating or a poor lipid profile that causes a large increase in near-term mortality risk, but no increase in longer-term mortality risk. The pattern is consistent with the former interpretation, e.g., a negative health shock between 2006 and 2014 causes both greater mortality risk in 2014-2018 and a decline in purpose between 2006-2014. In a companion study,^45^ we find evidence consistent with this interpretation as health shocks are strongly associated with a contemporaneous decline in purpose but a decline in purpose is largely independent of future health shocks.

Our analysis does not show purpose is unimportant (e.g., purpose in life likely increases happiness and life satisfaction) or has no casual impact on longevity. Rather, our evidence suggests that existing tests linking purpose levels to longevity suffer from reverse causation and if there is a causal link between purpose and longevity, it is much weaker than the estimates from previous studies suggest.

### Limitations

Our sample was limited to individuals 50 and older and purpose may better predict long-term longevity for a younger sample (although extant evidence suggests the reverse is true).^12^ Longer horizons also lead to more individuals lost to follow-up. Because most covariates are self-reported, there is also the possibility of common methods bias.

For tractability we focus solely on horizon and current health. Other variables (e.g., wealth, binge drinking) or variable specifications may confound the relation between purpose and mortality risk, i.e., our estimates likely still overestimate the relation between purpose and longevity. For example, a 79-year-old is likely to have both lower purpose and worse health than a 75-year-old (appendix p. 7), yet both are in the same age control group implying that variation in life purpose will capture within group age effects.

## Conclusions

The relation between life purpose and mortality is more tenuous than previous work suggests. Our evidence suggests a substantial portion of the relation between purpose and longevity arises because lower health causes lower purpose and higher mortality risk as the link between purpose and health quickly deteriorates with horizon or more thorough controls for current health levels. Although life purpose interventions may yield multiple benefits, the assertion that greater longevity will follow appears premature.

## Data Availability

All data were derived from publicly available Health and Retirement Study and detailed in the supplemental appendix.

https://hrs.isr.umich.edu/

## Contributors

This study was designed by RS and HT. Both authors accessed and verified the data. Both authors were involved with data interpretation and contributed to the methods development. RS did the data and statistical analyses. Both authors contributed to drafting the paper and revised the manuscript for important intellectual content. Both authors approved the decision to submit the final version of the manuscript.

## Data sharing

All data were derived from publicly available sources as detailed in the supplemental appendix.

## Acknowledgements

We are grateful for the support of the University of Arizona and Colorado State University.

