## Supplemental appendix for "Why does purpose in life predict mortality in older adults?"

**S1.** ***Supplementary Figure 1:*** **2006-2018 HRS cleaning flowchart**

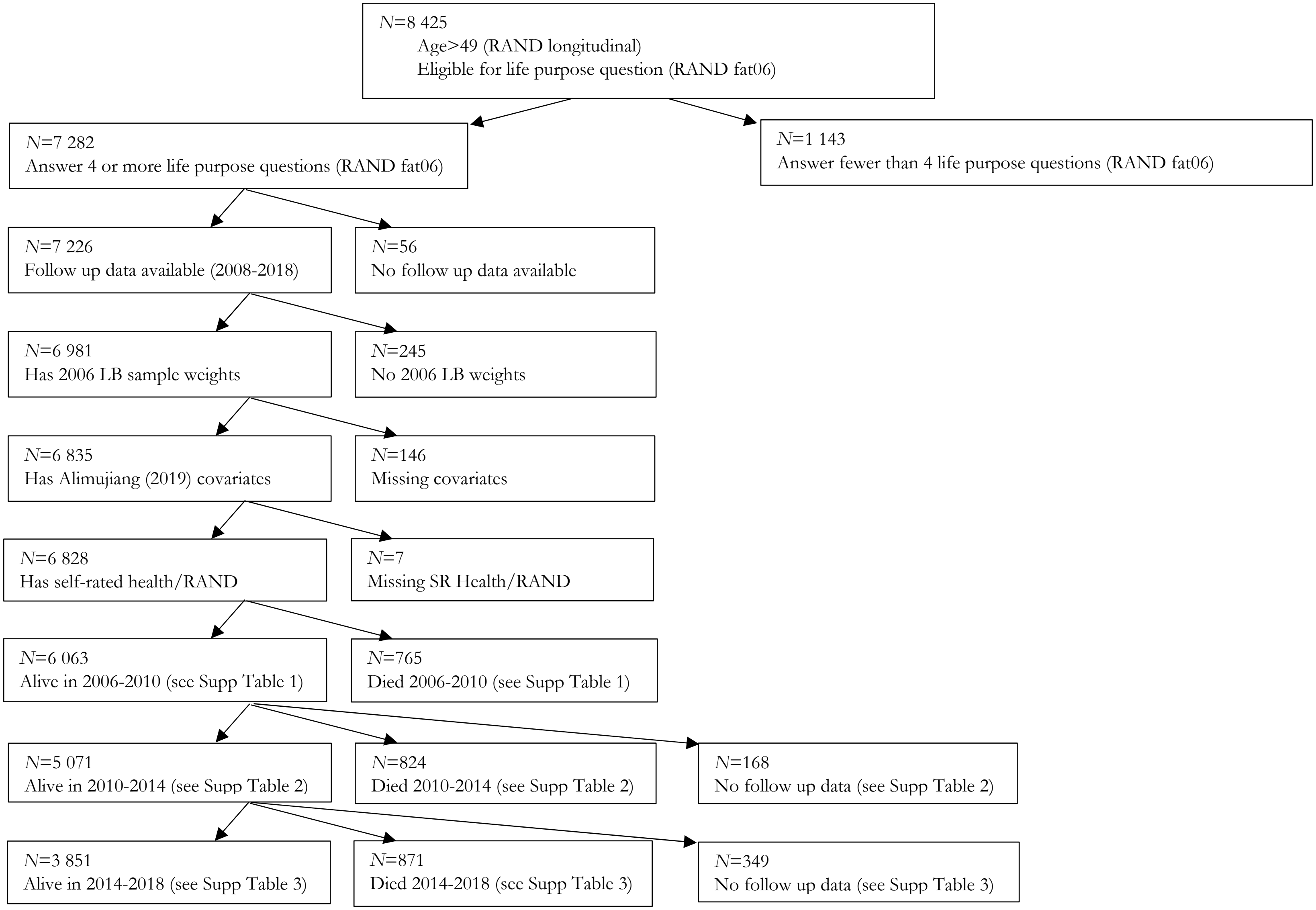

**S2.** ***Supplementary Figure 2:*** **2006-2018 Health and Retirement Study cleaning flowchart for Model 2**

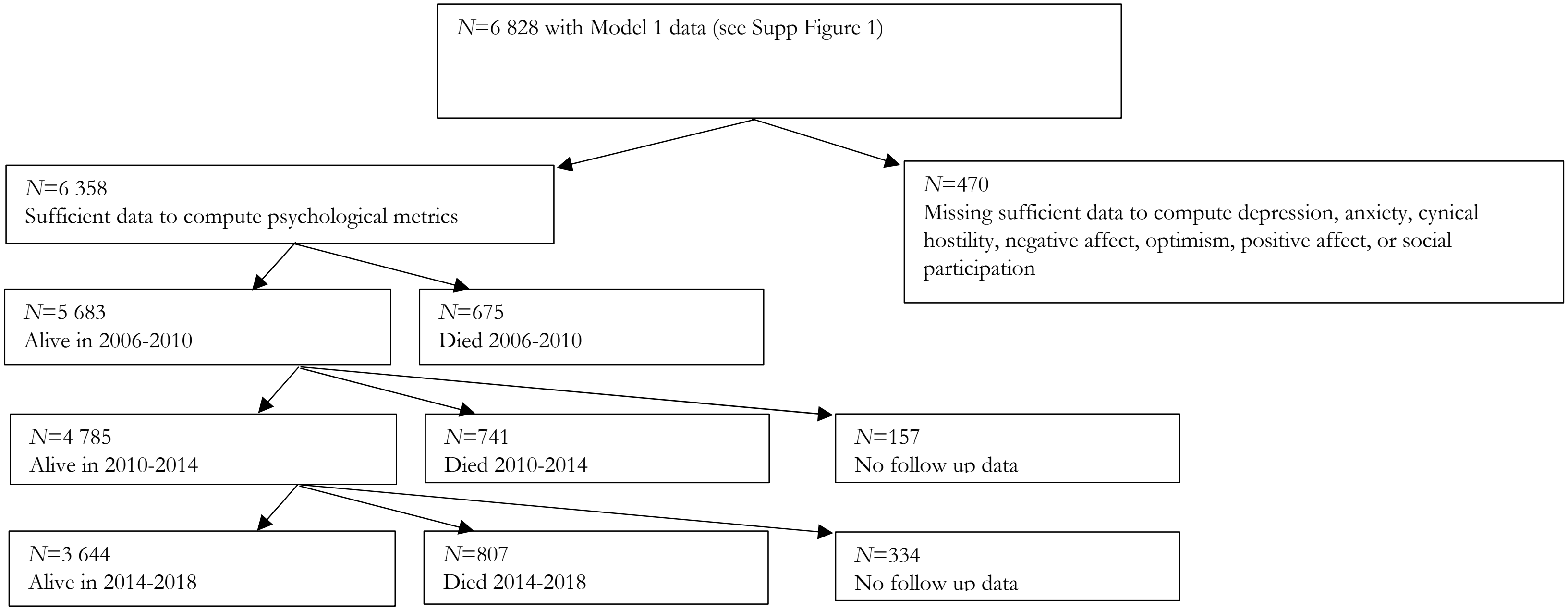

**S3. *Supplementary Table 1:* Censored and death 2006-2010**

| Dead or alive in 2010 | N | Case | Death/censored date |
|---|---|---|---|
| Dead 2006-2010 (1) | 765 | Reported died prior to 2010 interview | Reported death date |
| Alive (2) | 5 821 | Completed 2010 interview | Censored 2010 interview date |
| Alive (3) | 158 | Did not complete 2010 interview, but reported alive in subsequent (2012-2018) wave or reported died in a subsequent wave | Censored 48 months following 2006 interview date |
| Alive (4) | 84 | Did not complete 2010 interview or any subsequent interview. Did complete 2008 interview | Censored 2008 interview date |
| Total | 6 828 | (1) + (2) + (3) + (4) | |

### S4. *Supplementary Table 2:* Censored and death 2010-2014

| Dead or alive in 2014 | N | Case | Death/censored date |
|---|---|---|---|
| Died 2006-2010 (1) | 765 | | Died prior to 2010 interview |
| Died 2010-2014 (2) | 824 | Reported died with death date | Reported death date |
| Alive (3) | 4 813 | Interviewed in 2014 wave | Censored 2014 interview date |
| Alive (4) | 133 | Did not complete 2014 interview, but reported alive in subsequent (2016-2018) wave or reported died in a subsequent wave | Censored 96 months following 2006 interview date |
| Alive (5) | 125 | Last interviewed in 2012 wave | Censored 2012 interview date |
| Unknown (6) | 84 | Last interviewed in 2010 wave | Lacks follow up data |
| Unknown (7) | 84 | Last interviewed in 2008 wave | Lacks follow up data |
| 2010-2014 sample | 5 895 | (2) + (3) + (4) + (5) | |
| Total | 6 828 | (1) + (2) + (3) + (4) + (5) + (6) + (7) | |

### S5. *Supplementary Table 3:* Censored and death 2014-2018

| Dead or alive in 2018 | N | Case | Death/censored date |
|---|---|---|---|
| Died 2006-2010 (1) | 765 | | Died prior to 2014 interview |
| Died 2010-2014 (2) | 824 | | Died prior to 2014 interview |
| Died 2014-2018 (3) | 89 | Reported as died during 2018 wave, no death date given | Assumed death date=July 2017 |
| Died 2014-2018 (4) | 782 | Reported died with death date | Reported death date |
| Alive (5) | 3 433 | Interviewed in 2018 wave | Censored 2018 interview date |
| Alive (6) | 418 | Last interviewed in 2016 wave | Censored 2016 interview date |
| Unknown (7) | 224 | Last interviewed in 2014 wave | Lacks follow up data |
| Unknown (8) | 125 | Last interviewed in 2012 wave | Lacks follow up data |
| Unknown (9) | 84 | Last interviewed in 2010 wave | Lacks follow up data |
| Unknown (10) | 84 | Last interviewed in 2008 wave | Lacks follow up data |
| 2014-2018 sample | 4 722 | (3) + (4) + (5) + (6) | |
| Total | 6 828 | (1) + (2) + (3) + (4) + (5) + (6) + (7) + (8) + (9) + (10) | |

### S6. Differences in sample from Alimujiang et al.[1]

Our initial sample of individuals over age 49 and eligible for the 2006 life purpose questionnaire is slightly larger than that reported by Alimujiang et al.[1] (8 425 versus 8 419). This may arise because we use the RAND longitudinal file (which is more comprehensive) for the age calculation as well as updated HRS files. We are also close to the authors with respect to the number of respondents who answer four or more life purpose questions (N=7 282 versus 7 277). Because our data (we use the RAND longitudinal file) includes interviews up to the 2018 wave, we lose fewer observations to lack of follow up (56 versus 81 for Alimujiang et al.). In contrast, we lose more observations (242 versus 163 for Alimujiang et al.) due to missing (or zero) HRS weights. Approximately 11·2% of our sample of 6 828 respondents are reported to have died by 2010 interview versus 11·1% in the 6 985 respondents in the Alimujiang et al. sample.

Alimujiang[1] et al. report (in their supplemental material and in their study) that respondents must answer at least three of the seven life purpose questions to be included in the sample following guidance from HRS. The HRS guidance suggests respondents must answer at least four of the seven life purpose questions (i.e., "Set the final score to missing if there are more than three items with a missing value").[2] As a result, we require respondents answer at least four of the questions. Regardless, our figures closely align with those reported in Alimujiang et al.

As noted in the study, our descriptive statistics are nearly identical to those reported by the authors except for some variation in educational attainment. Specifically, we use the RAND definitions for educational level. RAEDUC identify less than high school, GED, high school graduate, some college, and college and above. RAND defines some college as more than 12 years of education and has a high school diploma or GED or if the degree is less than a BA or "other." RAEDEGRM is highest degree earned. We classify MA/MBA and Law/MD/PhD as graduate degrees.

### S7. *Supplementary Figure 3:* Age, life purpose, and self-rated health

As noted in the study and shown below (we plot the mean 2006 life purpose and self-rated health scores by age for ages with at least 25 observations), life purpose and self-rated health decline with age. The pooled cross-sectional correlation between age and life purpose is -0·17 (p<0·001, n=6 828) and between age and self-rated health is -0·13 (p<0·001, n=6 828). Thus, as older individuals die, the mean values increase (i.e., younger individuals are more likely to survive until 2014).

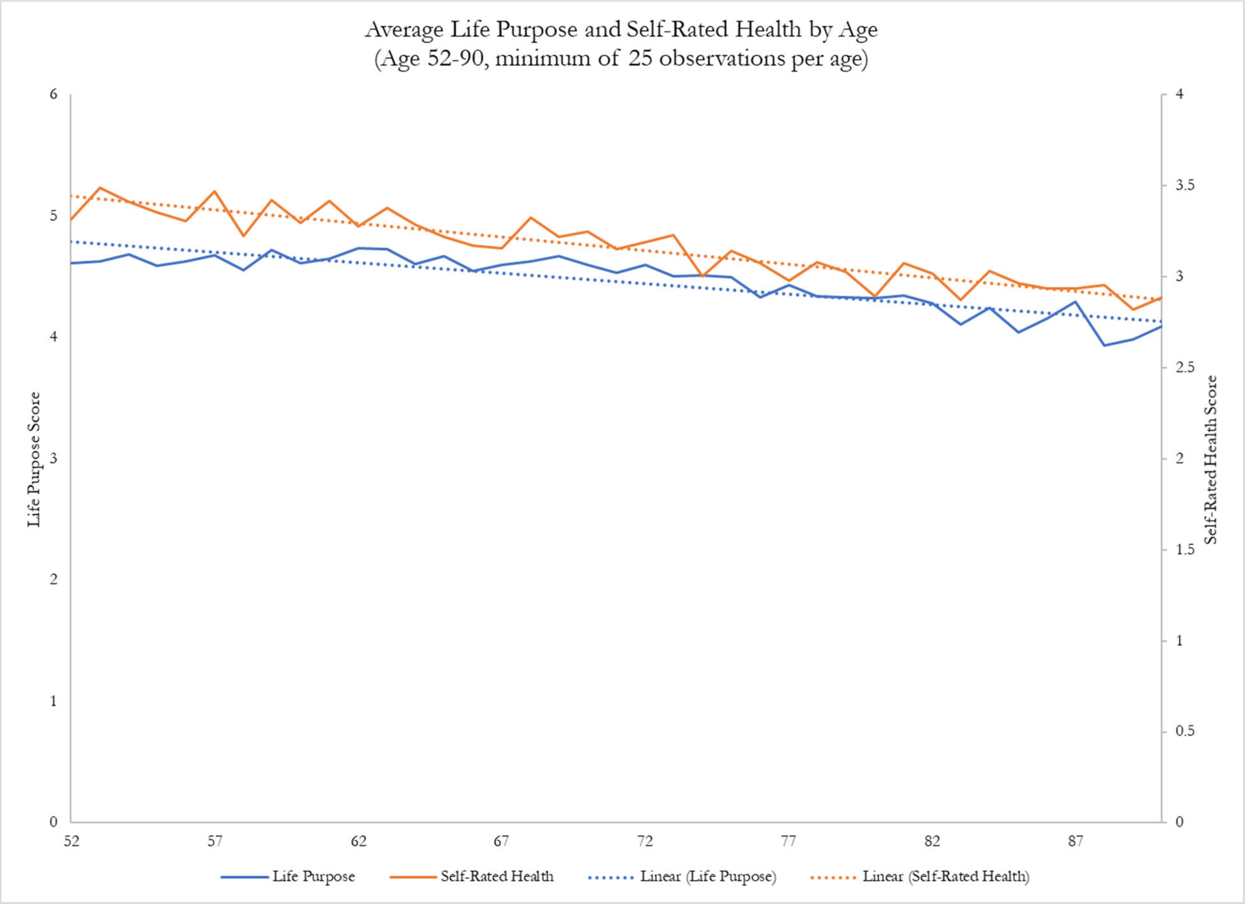

### S8. Additive survival/censoring times

To ensure that total survival times are additive (e.g., 2006-2018 survival=2006-2010 survival + 2010-2014 survival + 2018-2014 survival) we compute all values as differences from the 2006 interview date. For instance, for the second period (2010-2014) respondents with interviews in 2006, 2010, and 2014, the difference between the respondent's 2014 and 2010 survival times is the number of months between the 2010 interview and the 2014 interview. For respondents where we infer survival time (e.g., 96 months past their 2006 interview), the difference between the respondent's 2014 and 2010 survival times is 96 months less their 2010 interview date (for respondents with 2010 interviews) or 48 months (for respondents without 2010 interviews).

### S9. *Supplementary Table 4:* Variable definitions and sources

| Variable | Variable ID/construction | Source |
|---|---|---|
| Leave behind questionnaire weights | KLBWGTR | HRS Tracker file |
| Death date | RADDATE | RAND longitudinal file |
| Interview status | I8WBEG, I9WBEG, I10WBEG, I11WBEG, I12WBEG, I13WBEG, I14WBEG | RAND longitudinal file |
| Age (years) | R8AGEY_B | RAND longitudinal file |
| Sex | RAGENDER | RAND longitudinal file |
| Marital status | R8MSTAT<br>Married: R8MSTAT=1, 2, or 3<br>Divorced: R8MSTAT=4, 5, or 6<br>Widowed: R8MSTAT=7<br>Never married: R8MSTAT=8 | RAND longitudinal file |
| Race/ethnicity | RACACEM and RAHISPAN<br>White: RACACEM=1 and RAHISPAN=0<br>Black: RACEM=2<br>Hispanic White: RACACEM=1 RAHISPN=1<br>Other: RARACEM=3 | RAND longitudinal file |
| Education level | RAEDUC and RAEDEGRM<br>Less than high school: RAEDUC=1<br>High school graduate: RAEDUC=2 or RAEDUC=3<br>Some college: RAEDUC=4<br>College: RAEDUC=5 and RAEDEGRM NE 6 or 7<br>Graduate degree: RAEDEGRM=6 or 7<br>As noted in our study, we use the RAND definitions for educational level. RAEDUC identify less than high school, GED, high school graduate, some college, and college and above. RAND defines some college as more than 12 years of education and has a high school diploma or GED or if the degree is less than a BA or "other." RAEDEGRM is highest degree earned. We denote MA/MBA and Law/MD/PhD as graduate degrees. | RAND longitudinal file |
| Smoking status | R8SMOKEN and R8SMOKEV<br>Current smoker: R8SMOKEN=1<br>Former smoker: R8SMOKEN NE 1 and R8SMOKEV=1<br>Never smoker: R8SMOKEV=0 | RAND longitudinal file |
| Alcohol consumption | R8DRINKD<br>0: R8DRINKD=0<br>1-2: R8DRINKD=1 or R8DRINKD=2<br>3-4: R8DRINKD=3 or R8DRINKD=4<br>5-6: R8DRINKD=5 or R8DRINKD=6<br>7: R8DRINKD=7 | RAND longitudinal file |
| BMI | R8BMI<br>Low BMI: R8BMI<18·5<br>Normal BMI: 18·5 ≤ R8BMI<25<br>Overweight BMI: 25 ≤ R8BMI<30<br>Obese BMI: R8BMI>30 | RAND longitudinal file |

| Variable | Variable ID/Construction | Source |
|---|---|---|
| Vigorous physical exercise | R8VGACTX<br>Daily: R8VGACTX=1<br>>1/week: R8VGACTX=2<br>1/week: R8VGACTX=3<br>1-3/month: R8VGACTX=4<br>Hardly ever or never: R8VGACTX=5 | RAND longitudinal file |
| High blood pressure | KC005=1 | RAND 2006 fat file |
| Diabetes | KC010=1 | RAND 2006 fat file |
| Cancer | KC018=1 | RAND 2006 fat file |
| Lung disease | KC030=1 | RAND 2006 fat file |
| Heart disease | KC036=1 | RAND 2006 fat file |
| Stroke | KC053=1 | RAND 2006 fat file |
| Chronic illness | If any of the above (KC005, KC010, KC08, KC030, KC036, or KC053), equals 1, chronic disease=1 | RAND 2006 fat file |
| Functional score | R8ADLA | RAND longitudinal file |
| Life purpose score | For individuals who answer at least four of the seven questions: LP=sum(KLB035A,-1*KLB035B,KLB035C,-1*KLB035D, -1*KLB035E,-1*KLB035F,KLB035G) | RAND 2006 fat file |
| Self-rate health | R8SHLT<br>Self-rated health=6-R8SHLT | RAND longitudinal file |
| Broad limitations | First principal component of R8MOBILA, R8LGMUSA, R8GROSSA, and R8FINEA | RAND longitudinal file |
| | Additional model 2 covariates (construction of these variables follows HRS recommendations)[2] | |
| Anxiety | For individuals who answer at least three of the following: Anxiety=average of (KLB041A,KLB041B, KLB041C,KLB041D,KLB041E) | RAND 2006 fat file |
| Cynical hostility | For individuals who answer at least two of the following: Cynical hostility=average of (KLB019A,KLB019B,KLB019C, KLB019D,KLB019) | RAND 2006 fat file |
| Negative affect | For individuals who answer at least three of the following: Negative affect=6+mean(-1* KLB027I,-1*KLB027J, -1*KLB027K,-1*KLB027L,-1*KLB027M,-1*KLB027N) | RAND 2006 fat file |
| Optimism | For individuals who answer at least four of the following: Optimism=average of (-1*KLB019F,-1*KLB019J, -1* KLB019K,KLB019G,KLB019H,KLB019I) | RAND 2006 fat file |
| Positive affect | For individuals who answer at least three of the following: Positive affect=6 + average of (-1* KLB027A,-1*KLB027B, -1*KLB027C,-1*KLB027D,-1*KLB027E,-1*KLB027F) | RAND 2006 fat file |
| Social participation | For individuals who answer at least four of the following: Social participation=sum(SP1,SP2,SP3,SP4,SP5,SP6,SP7).<br>SP1=1 if KLB001A=1, SP2=1 if KLB001B=1,<br>SP3=1 if KLB001C=1, SP4=1 if KLB001D=1,<br>SP5=1 if KLB001E=1, SP6=1 if KLB001F=1,<br>SP7=1 if KLB001G=1 | RAND 2006 fat file |
| Depression | R8CESD for observations with no missing values (i.e., R8CESDM=0) | RAND longitudinal file |

### S10. *Supplementary Table 5:* Hazard ratios for individual chronic diseases from Model 1A

| | Hazard ratio (95% CI) | | |
|---|---|---|---|
| **Chronic illness (2006)** | **2006-2010** | **2010-2014** | **2014-2018** |
| High blood pressure | 1·08 (0·91-1·28) | 0·91 (0·77-1·08) | 1·13 (0·95-1·33) |
| Diabetes | 1·49 (1·24-1·78)[a] | 1·40 (1·17-1·67)[a] | 1·22 (1·01-1·47)[b] |
| Cancer | 1·19 (0·99-1·42) | 1·27 (1·05-1·52)[b] | 1·23 (1·01-1·50)[b] |
| Lung disease | 1·36 (1·12-1·66)[b] | 1·41 (1·14-1·74)[b] | 1·04 (0·82-1·33) |
| Heat disease | 1·30 (1·10-1·54)[b] | 1·20 (1·02-1·42)[b] | 1·26 (1·06-1·49)[b] |
| Stroke | 1·03 (0·81-1·31) | 0·99 (0·75-1·29) | 1·40 (1·07-1·83)[b] |

We report Cox proportional hazard model results adjusting for age, sex, education, marital status, smoking status, frequency of vigorous physical activity, days/week consuming alcohol, body mass index, functional status, indicators for diagnosis of hypertension, diabetes, cancer, lung disease, heart disease, and stroke, and self-rated health with 2006 weights. [a] $p$<0·001. [b] $p$<0·05.

### S11. Broad limitations calculation

We compute broad limitations as the first principal component of the four RAND functional limitation metrics: Mobility (RwMOBILA), Large Muscle (RwLGMUSA), Gross Motor Skills (RwGROSSA), and Fine Motor Skills (RwFINEA). Each index is a sum of the number of functions that the respondent has "some difficulty." Mobility includes walking one block, walking several blocks, walking across a room, climbing one flight of stairs, and climbing several flights of stairs. Large muscle includes sitting for two hours, getting up from a chair, stooping, kneeling or crouching, and pushing or pulling large objects activities. The gross motor skills include walking one block, walking across a room, climbing one flight of stairs, getting in or out of bed, and bathing activities. Fine motor skills include picking up a dime, eating, and dressing. The first principal component explains 68% of the total variation in the four factors and has an eigenvalue of 2·706.

### S12. *Supplementary Table 6:* Constant proportionality tests

The Cox proportional hazard model assumes that life purpose has a constant impact on mortality risk. To examine this assumption, we follow the recommendation in the literature[3,4] and add a variable interacting life purpose with the natural logarithm of time. Specifically, we estimate the baseline (i.e., Model 1) model but replace the life purpose group indicator variables with the individual's life purpose score and the product of the individual's life purpose score and the natural logarithm of 1+number of months (until censor or death). A statistically significant value of the interaction term indicates the relation between life purpose and mortality risk varies over time. Because we interact the natural logarithm of time with life purpose score (i.e., a higher value indicates greater life purpose), a hazard ratio greater than one for the interaction term indicates the relation between purpose and mortality risk declines with time.

| | **Hazard ratio (95% CI)** | |
|---|---|---|
| | **2006-2010** | **2006-2018** |
| Life purpose score | 0·51 (0·36-0·71)[a] | 0·56 (0·45-0·71)[a] |
| Life purpose*ln(1+no. months) | 1·15 (1·04-1·28)[b] | 1·11 (1·05-1·17)[a] |

Cox proportional hazard model analysis adjusting for age, sex, education, marital status, smoking status, frequency of vigorous physical activity, days/week consuming alcohol, body mass index, functional status, an indicator for the presence of a chronic condition, life purpose score and an interaction term computed as the product of life purpose score and (1+ number of months until censored or death) with 2006 weights. [a] $p<0{\cdot}001$. [b] $p<0{\cdot}05$.

**S13.** ***Supplementary Figure 4:*** **2014-2018 Health and Retirement Study cleaning flowchart**

This figure reports the data cleaning process of the 2014-2018 period for the analysis considered in "Discussion" section. This sample includes all individuals in the 2014 HRS dataset over the age of 49 and eligible for the HRS 2014 life purpose questions. Supplementary Table 7 estimates a Cox survival model for the 2014-2018 period based on this sample. Supplementary Table 8 estimates a Cox survival model for the 2014-2018 period when limiting the sample to respondents with both 2006 life purpose and covariate data and 2014 life purpose and covariate data.

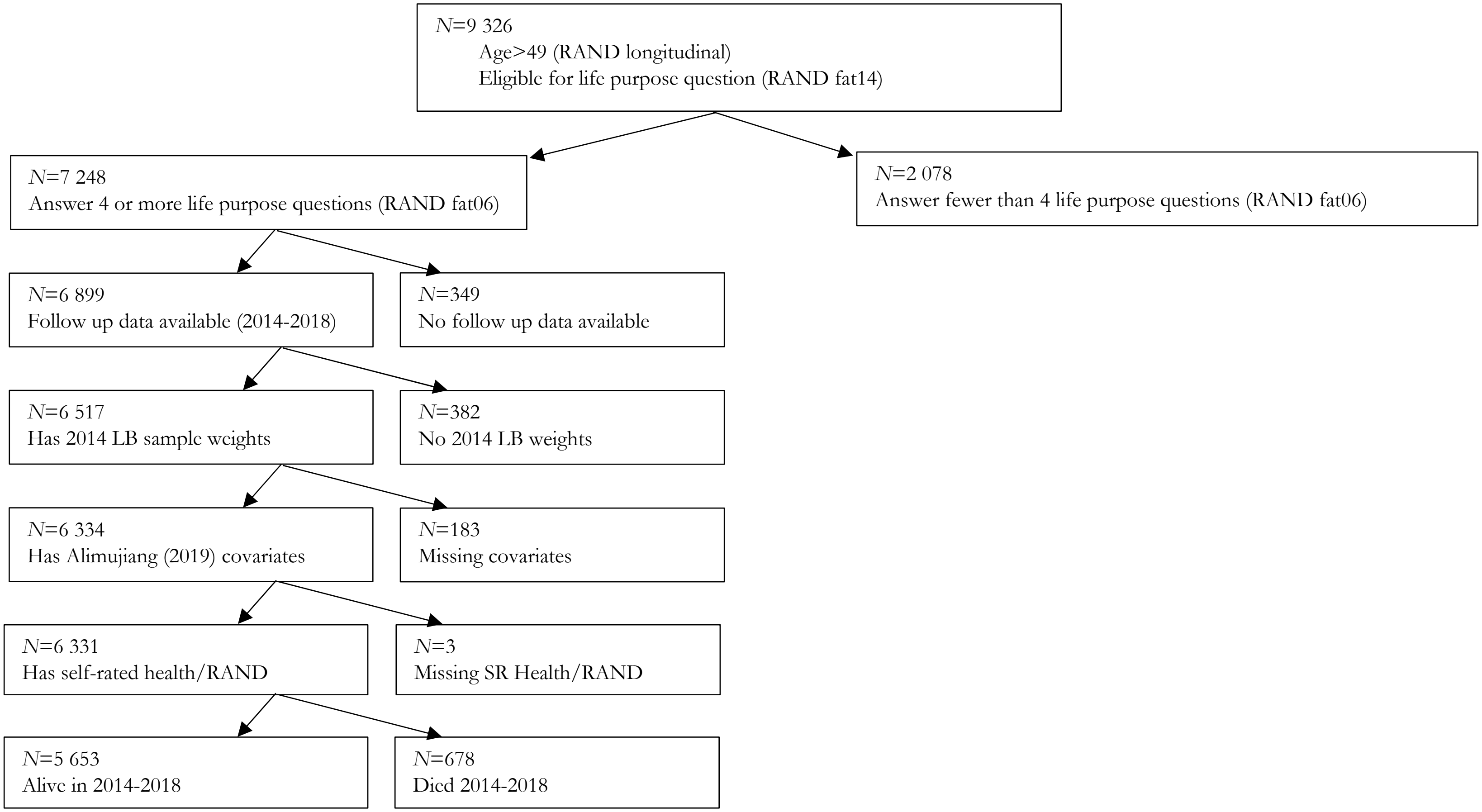

**S14.** ***Supplementary Table 7:*** **Life purpose, health, and all-cause mortality 2014-2018**

| Life purpose score category | Hazard ratio (95% CI) | | |
|---|---|---|---|
| | **Model 1** | **Model 1A** | **Model 1B** |
| 2014-2018, n=6 331 Health and Retirement Study participants | | | |
| 1·00-2·99 | 2·64 (1·51-4·64)[a] | 1·95 (1·11-3·44)[b] | 2·16 (1·23-3·80)[b] |
| 3·00-3·99 | 1·93 (1·15-3·26)[b] | 1·56 (0·92-2·64) | 1·62 (0·96-2·74)[b] |
| 4·00-4·99 | 1·31 (0·78-2·22) | 1·16 (0·69-1·96) | 1·17 (0·69-1·98) |
| 5·00-5·99 | 1·09 (0·64-1·85) | 1·01 (0·59-1·73) | 1·01 (0·59-1·73) |
| 6·00 | 1 [Reference] | 1 [Reference] | 1 [Reference] |
| Chronic/individual disorders | Chronic indicator | Individual disorders | Individual disorders |
| Functional score | 1·31 (1·23-1·40)[a] | 1·21 (1·13-1·29)[a] | |
| Self-rated health | | 0·75 (0·68-0·83)[a] | |
| Broad limitations | | | 1·39 (1·30-1·49)[a] |

Model 1 reports Cox proportional hazard model for the 2014-2018 period based on life purpose and other covariates measured in 2014. The sample (see Supplementary Figure 4) consists of the 6 331 individuals with sufficient data in 2014. Covariates include age, sex, education, marital status, smoking status, frequency of vigorous physical activity, days/week consuming alcohol, body mass index, functional status, and an indicator for the presence of a chronic condition. Model 1A reports Cox proportional hazard model results analogous to Model 1 except Model 1A also includes self-rated health as a regressor, and the indicator of a chronic condition is replaced by individual disorder indicators for diagnosis of hypertension, diabetes, cancer, lung disease, heart disease, and stroke. Model 1B is analogous to Model 1A except the functional score and self-rated health regressors are replaced with the first principal component (denoted broad limitations) from the RAND measures of Mobility Problems, Gross Motor Problems, Large Muscle Problems, and Fine Motor Problems a broad limitations measure. All Cox proportional hazard models were weighted by 2014 weights. [a] $p$<0·001. [b] $p$<0·05.

### S15. ***Supplementary Table 8:*** 2014-2018 analysis limited to respondents with 2006 and 2014 data

We limit the sample to individuals who have both 2006 life purpose and covariate data and 2014 life purpose and covariate data. We then estimate Cox survival models for the four-year period 2014-2018 on 2006 life purpose and covariates (top panel) and 2014 life purpose and covariates (bottom panel).

| | Hazard ratio (95% CI) | | |
|---|---|---|---|
| **Life purpose score category** | **Model 1** | **Model 1A** | **Model 1B** |
| 2014-2018, n=3 928 HRS participants 2006 life purpose and covariates[a] | | | |
| 1·00-2·99 | 0·90 (0·48-1·69) | 0·80 (0·42-1·51) | 0·85 (0·45-1·6) |
| 3·00-3·99 | 1·18 (0·76-1·83) | 1·07 (0·69-1·66) | 1·12 (0·72-1·75) |
| 4·00-4·99 | 0·94 (0·61-1·44) | 0·86 (0·56-1·33) | 0·90 (0·59-1·39) |
| 5·00-5·99 | 0·77 (0·50-1·20) | 0·76 (0·49-1·19) | 0·79 (0·50-1·22) |
| 6·00 | 1 [Reference] | 1 [Reference] | 1 [Reference] |
| Chronic/individual disorders | Chronic indicator | Individual disorders | Individual disorders |
| Functional score | 1·53 (1·19-1·97)[c] | 1·10 (0·97-1·25) | |
| Self-rated health | | 0·76 (0·68-0·85)[c] | |
| Broad limitations | | | 1·21 (1·08-1·34)[c] |
| 2014-2018, n=3 928 HRS participant 2014 life purpose and covariates[b] | | | |
| 1·00-2·99 | 1·94 (1·23-3·07)[d] | 1·52 (0·96-2·41) | 1·70 (1·07-2·68)[d] |
| 3·00-3·99 | 1·51 (1·02-2·23)[d] | 1·29 (0·87-1·91) | 1·32 (0·89-1·95) |
| 4·00-4·99 | 1·03 (0·69-1·53) | 0·94 (0·63-1·41) | 0·94 (0·63-1·41) |
| 5·00-5·99 | 0·76 (0·5-1·16) | 0·75 (0·49-1·15) | 0·73 (0·48-1·12) |
| 6·00 | 1 [Reference] | 1 [Reference] | 1 [Reference] |
| Chronic/individual disorders | Chronic indicator | Individual disorders | Individual disorders |
| Functional score | 1·96 (1·37-2·8)[c] | 1·23 (1·14-1·33)[c] | |
| Self-rated health | | 0·77 (0·70-0·86)[c] | |
| Broad limitations | | | 1·44 (1·32-1·58)[c] |

Model 1 reports Cox proportional hazard model adjusting for age, sex, education, marital status, smoking status, frequency of vigorous physical activity, days/week consuming alcohol, body mass index, functional status, and an indicator for the presence of a chronic condition. Model 1A reports Cox proportional hazard model results analogous to Model 1 except Model 1A also includes self-rated health as a regressor, and the indicator of a chronic condition is replaced by individual disorder indicators for diagnosis of hypertension, diabetes, cancer, lung disease, heart disease, and stroke. Model 1B is analogous to Model 1A except the functional score and self-rated health regressors are replaced with the first principal component (denoted broad limitations) from the RAND measures of Mobility Problems, Gross Motor Problems, Large Muscle Problems, and Fine Motor Problems a broad limitations measure. [a] Life purpose, covariates, and weights based on 2006 data. [b] Life purpose, covariates, and weights based on 2014 data. [c] $p<0·001$. [d] $p<0·05$.

### S16. STROBE statement

Following Alijumiang et al,[1] and for the sake of brevity, the present study followed the Strengthening the Reporting of Observational Studies in Epidemiology (STROBE) reporting guideline for cohort studies with the exception of reporting multiple levels of confounder-adjusted estimates (rather than including fully unadjusted estimates).
